# COVID-19 control across urban-rural gradients

**DOI:** 10.1101/2020.09.07.20189597

**Authors:** Konstans Wells, Miguel Lurgi, Brendan Collins, Biagio Lucini, Rowland R. Kao, Alun L. Lloyd, Simon D.W. Frost, Mike B. Gravenor

## Abstract

Controlling the regional re-emergence of SARS-CoV-2 after its initial spread in ever-changing personal contact networks and disease landscapes is a challenging task. In a landscape context, contact opportunities within and between populations are changing rapidly as lockdown measures are relaxed and a number of social activities re-activated. Using an individual-based metapopulation model, we explored the efficacy of different control strategies across an urban-rural gradient in Wales, UK. Our model shows that isolation of symptomatic cases, or regional lockdowns in response to local outbreaks, have limited efficacy unless the overall transmission rate is kept persistently low. Additional isolation of non-symptomatic infected individuals, who may be detected by effective test and trace strategies, is pivotal to reduce the overall epidemic size over a wider range of transmission scenarios. We define an ‘urban-rural gradient in epidemic size’ as a correlation between regional epidemic size and connectivity within the region, with more highly connected urban populations experiencing relatively larger outbreaks. For interventions focused on regional lockdowns, the strength of such gradients in epidemic size increased with higher travel frequencies, indicating a reduced efficacy of the control measure in the urban regions under these conditions. When both non-symptomatic and symptomatic individuals are isolated or regional lockdown strategies are enforced, we further found the strongest urban-rural epidemic gradients at high transmission rates. This effect was reversed for strategies targeted at symptomatics only. Our results emphasise the importance of test-and-tracing strategies and maintaining low transmission rates for efficiently controlling COVID19 spread, both at landscape scale and in urban areas.

**Author summary:** The spread of infectious diseases is the outcome of contact patterns and involves source-sink dynamics of how infectious individuals spread the disease through pools of susceptible individuals. Control strategies that aim to reduce disease spread often need to accept ongoing transmission chains and therefore, may not work equally well in different scenarios of how individuals and populations are connected to each other. To understand the efficacy of different control strategies to contain the spread of COVID19 across gradients of urban and rural populations, we simulated a large range of different control strategies in response to regional COVID19 outbreaks, involving regional lockdown and the isolation individuals that express symptoms and those that developed not symptoms but may contribute to disease transmission. Our results suggest that isolation of asymptomatic individuals through intensive test-and-tracing is important for efficiently reducing the epidemic size. Regional lockdowns and the isolation of symptomatic cases only are of limited efficacy for reducing the epidemic size, unless overall transmission rate is kept persistently low. Moreover, we found high overall transmission rates to result in relatively larger epidemics in urban than in rural communities for these control strategies, emphasising the importance of keeping transmission rates constantly low in addition to regional measures to avoid the disease spread at large scale.

## Introduction

In the absence of a vaccine against COVID-19 during the initial pandemic phase, stakeholders are confronted with challenging decision-making to balance constraints of social interaction and the efficient isolation of infectious individuals with economic and social pressures. There is now growing scientific evidence of how different containment strategies compare to each other amid the challenges of asymptomatic disease transmission and the ongoing need for improved estimates of epidemiological key parameters [1, 2]. Non-pharmaceutical interventions for curbing the spread of COVID-19 rely on the isolation of infectious individuals or general social distancing policies to reduce interactions between undetected infectious individuals and those susceptible to the disease. During uncontrolled pandemic spread, a central aim is to reduce case incidence in order to release the pressure on health systems. A more fundamental, long-term, goal should be to reduce the overall epidemic size and allow particularly those most prone to suffer from the disease to escape infection until a pharmaceutical measure such as a vaccine is in place.

Control strategies are likely to be regional, and temporal, aiming to reduce the time-dependent reproduction number *R*, while accepting that ongoing transmission is long term. But how should these regional and temporary strategies account for disease spread in ever-changing transmission landscapes? One particular question faced by many countries is how do different control strategies differ in their efficacy in preventing disease spread across urban-rural gradients of different population densities and connectivity in urban and rural landscapes?

The spread of infectious disease is rarely random. It is instead likely driven by the complex and heterogeneous social interaction patterns of humans and the stark gradient between urban and rural populations. In a landscape context, contact opportunities within and among populations across urban-rural gradients, and source-sink dynamics arising from infectious individuals encountering pools of susceptible individuals, are the ultimate drivers of disease spread. Disease spread is thus hampered if contact opportunities are lower in poorly mixed populations [3–5]. Heterogeneity in contact patters of individuals and among social groups is also assumed to impact the depletion of the pool of susceptible individuals and the build-up of possible herd immunity that prevent further spread [6, 7]. Hence, future short- and long-term mitigation strategies that focus on managing regional and erratic outbreaks would benefit from a better understanding of which control strategies provide the best possible outcome under variable regional conditions.

To the best of our knowledge, there is so far little evidence of how various disease control strategies differ in their efficacy across urban-rural gradients [8]. To address this gap, using an individual-based metapopulation model, we explore the outcomes of different control strategies to contain the epidemic size of COVID-19 in ever changing disease landscapes of case numbers and susceptible depletion, which involve strong urban-rural gradients.

Our modelling approach is strategic, in contrast to many tactical COVID-19 simulation models that have focused on replication of specific characteristics of real outbreaks with the aim of predicting the epidemic in specific locations [1, 9, 10]. Rather than modelling a certain scenario, we aim to define wide ranges and explore the model behaviour across a large array of combinations of transmission and control parameters. The influence of each parameter on particular outcomes can then be explored statistically. In this manner we aim to highlight how basic properties of realistic metapopulations’ structure that include urban-rural gradients, can affect the impact of control measures.

## Methods

### Case study of a rural-urban metapopulation in Wales

In order provide an empirical basis to explore possible COVID-19 spread across an urban-rural gradient and the efficacy of different disease control measures, we selected four counties in southwestern Wales (Pembrokeshire, Carmarthenshire, Swansea, Neath Port Talbot) with a total human population size of 701,995 (hereafter termed ‘metapopulation’) dispersed over an area of 4,811 km^2^ as a case study. This area was selected because of its strong urban-rural gradient, from city centres to sparsely occupied farming localities, and readily available demographic data.

We used demographic data from the United Kingdom 2011 census (Office for National Statistics, 2011, http://www.ons.gov.uk), and constructed a metapopulation model at the level of Lower Layer Super Output Area (LSOA), which provided *M* = 422 geographical units of regional populations with a mean of 1,663 individuals (SD = 387) each.

We used a gravity model to define the connections between populations, as it is capable of reflecting the connectivity underpinning landscape-scale epidemics [11, 12]. In particular, a gravity model was chosen as the LSOA administrative units are characterized by fairly similar population sizes, although they can have widely areas and hence different population densities. We calculated for each pair of populations a gravity measure *T_i,j_* of the relative strength of how individuals are attracted to population *i* from populations *j* by accounting for local population sizes *N* and weighted pairwise Euclidian distance measures *d^ζ^*, including the ten nearest populations *k* of the attractive population:

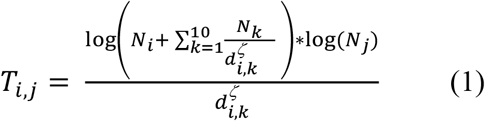

We assumed this approach to reflect reasonably well situations in which people are most attracted to higher density population clusters of urban populations (i.e. Swansea in our case study; the arbitrary selected number of ten nearest populations generates larger values of *T_i,j_* if the attractant population is closely surrounded by others; **Fig S1**). The scaling factor ζ (0 ≤ ζ ≤ 1) is a sampled parameter that may vary across scenarios, accounting for the uncertainty in population connectivity. For each population *i*, we computed a regional gravity index (with self-terms of 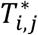 for *i* = *j* being zero):

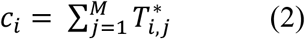

based on the scaled (mean subtracted from values divided by 1 SD) values of *T_i,j_* (denoted 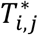), which we assumed to reflect the overall connectivity of the population within the global metapopulation. We used values of 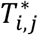 multiplied by the commuter travel frequency among populations (*ρ*) to compute the number of individuals visiting each population from elsewhere.

Within each local patch in the metapopulation, individuals encounter each other depending on their social interactions. The daily within-population contact numbers *F_i,t_* for any individual *i* at time *t* is assumed to be a random draw given by the sum of contacts drawn from a negative binomial (with r = 3 and p = 0.26, resulting in contact numbers with mean of 9 and SD of 6) and lognormal distribution (with mean = 3 and SD = 2, resulting in additional contact numbers with mean of 12 and SD of 16), whereby the lognormal distribution accounts for the ‘long-tail’ of contact frequency distributions. These parameters were based on a previous study of social contact frequencies in the UK [13]. For simplicity, and having in mind the main focus of this study on metapopulation-level patterns of disease spread, we did not account for repeated contact with the same individuals such as household or group members over different days. For simplicity, commuting individuals were assumed to return to their home populations in each time step, and their contacts were draw in the same way as for non-commuting individuals.

### Modelling the outcome of different disease control strategies in variable disease landscapes

We ran numerical simulations of an individual-based stochastic difference equation S-E-A-I-R model at daily time steps (see **Supplementary materials**), with individuals transitioning from a (S)usceptible compartment to being (E)xposed if infected. Exposed individuals become either infectious and symptomatic (I) or infectious but asymptomatic (A) after an incubation period of *τ* days. They then transition to a (R)emoved compartment with the recovery rate *γ*, which removes them from taking any further part in the transmission cycle. Both symptomatic and asymptomatic individuals can expose those susceptible to the virus.

The force of infection *λ_i,t_*, i.e. the probability that a susceptible individual *i* acquires SAR-SCoV-2 at time *t*, is calculated by considering the probabilities of the virus being transmitted from any interacting infected individual *k* (with *k* ∈ 1…*K_i,t_*, and *K_i,t_* being the number of all infectious individuals in the randomly sampled daily contact number *F_i,t_* of individual *i*); *λ_i,t_* can be computed based on the probability that none of the contact events with an infectious individual leads to an infection:

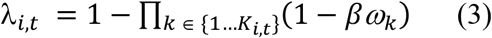

where *β* is the disease transmission parameter, and *ω_k_* is a scaling factor of infectiousness of asymptomatic relative to infectious individuals with 0 < *λ_i,t_* < 1.

To explore different scenarios of local and global epidemic sizes, we accounted for different pandemic stages and uncertainty in epidemiological parameters by varying systematically the following six parameters (see Supplementary Material, Table S1):

1. Transmission parameter (*β*),
2. The proportion of individuals that remain asymptomatic after infection (φ),
3. The relative infectiousness of asymptomatic disease carriers (ω),
4. Commuter travel frequency of individuals between populations (*ρ*),
5. Density dependence of individual contact numbers (*δ*),
6. Proportion of the overall population resistant/ recovered from infection at the onset of simulations.

Density-dependence of contact numbers (a population-level attribute) was modelled by calculating the scaled regional population density (i.e. all values divided by maximum density) to the power of the parameter *δ* and multiplied the corresponding values with the lognormal (‘long-tail’) component of the daily contact numbers *F_i,t_*. The resulting value corresponds to the same contact frequencies if *δ* approaches zero and truncated contact frequencies at low population densities if *δ* approaches one. Due to the lack of better empirical evidence, we assumed this approach to represent the situation in which an increase in population density (in urban areas) can result in a larger overall number of random encounters between citizens and higher contact frequencies between individuals of the same community in urban areas [14].

To assess and compare the efficacy of different, idealized, disease control strategies, we defined three general control strategies:

i. Trace and isolation of any infected individuals with a certain proportion (*κ*) of all infected individuals successfully isolated (removal of individuals in disease states E, A, I, reflecting scenarios where intensive and continuous testing and/or intensive contact tracing would allow removal of any infected individuals; termed ‘trace all’ in figures).
ii. Trace and isolation of symptomatic individuals only with a certain proportion (*ε*) of symptomatic individuals successfully isolated (removal of individuals in disease state I, reflecting scenarios where symptomatic cases isolate without any additional contract tracing or testing; termed ‘trace symptomatic only’ in figures).
iii. Regional temporary reduction of transmission rates (‘regional lockdown’) in response to a regional outbreak within the modelled LSOA administrative units, with four parameters to vary for decision making and control: (1) a threshold *α* defining the proportion of the regional population to be in disease state I, (2) lockdown stringency *ϕ* (the factor by which the transmission parameter is reduced), (3) travel ban distance *ν* (the maximum distance from which individuals are allowed to visit a locked-down population), and (4) duration of regional lockdown (*η*).

For simplicity, we did not account for possible individual heterogeneity in transition probabilities between different disease states but rather assumed constant ‘average’ transition probabilities in each scenario, albeit waiting times at different disease states are heterogeneous for many infectious diseases [15]. Similarly, we assume that the delay in the detection of individuals in different disease states is covered in the ‘average’ parameter of tracing/removing these individuals from transmission cycles as part of control strategies. We do so as here we are solely interested in population level outcomes of COVID19 spread in response to different control strategies.

### Numerical simulations

To be able to assess the efficacy of these control strategies as compared to a reference, we defined 10,000 ‘baseline’ transmission scenarios by varying the epidemiological parameters defining the spread scenarios (1–6 above). We performed independent numerical simulations for each parameter combination. We then combined each baseline transmission scenario with varying parameters for each of the three control strategies, running a total of 40,000 simulations, each for a time period of 100 days, which we assumed to be sufficiently long to capture the epidemic dynamics in response to different parameter values. Parameter values were sampled using latin hypercube sampling [16]; see Table S1 for ranges of parameter values used.

We started each simulation by randomly allocating n = 422 individuals as infectious (corresponding to the number of populations, but not necessarily one infectious individual in each population and infectious individuals are not necessarily seeded in high density populations) in the metapopulation. While this seeding of the epidemic does not represent any particular ‘true’ epidemic state in the studied population, we have chosen this the seeding together with the varying number of initially resistant proportion of populations to enable us to explore different scenarios of dynamic disease landscapes rather than any particular past or current state.

### Output summary

For each simulation, we computed the epidemic sizes as the numbers of individuals that had been symptomatic (we considered symptomatic cases only as asymptomatic cases are less likely to result in hospitalization or any other severe health burden) for each population and at the metapopulation scale (i.e. entire population). In order to explore the sensitivity of different control strategies to different epidemiological parameters, we calculated the relative differences in epidemic sizes (‘relative epidemic size’) for each disease control scenario and the corresponding baseline scenario at regional and metapopulation scale such that values close to zero mean effective control and larger values mean less effective control. Moreover, we computed for each baseline scenario the strength of correlation (expressed as the *r* value from Spearman rank correlation) between the regional relative epidemic size and the respective regional gravity index (‘urban-rural gradient in relative epidemic size’) in order to explore whether control strategies varied in their efficacy across urban-rural gradients. A strong positive correlation can be interpreted as a strong urban-rural gradient of disease spread, with smaller relative epidemic sizes in rural areas, where connectivity is generally lower. We also computed the strength of correlation between the epidemic sizes of baseline scenarios (uncontrolled outbreaks) and the respective regional gravity index.

In order to explore variation in the relative epidemic size and efficacy of different control strategies for different scenarios, we used generalised linear models (GLMs) and boosted regression trees (BRT) as implemented in the R package *dismo* [17]. We express results in terms of direction of effects (i.e. decrease/increase in relative epidemic size, reflecting higher/lower control efficacy) and relative influence (i.e. % of variance explained by various parameters in the corresponding BRT model) for those parameters that appear to show ‘significant’ effects in both GLM and BRT (i.e. GLM coefficients clearly distinct from zero, relative parameter influence > 5%).

All analyses and plotting were conducted in R version 4.0 [18].

## Results

The urban-rural gradient in epidemic sizes (expressed as rank correlation coefficient between the regional epidemic size and the regional gravity index) considerably decreased among baseline scenarios (uncontrolled outbreaks) with larger transmission parameters (β, explaining 57% of changes in total epidemic sizes). This indicates that larger outbreaks in urban areas occur mostly at low transmission parameters. In addition, the urban-rural gradient in total epidemic sizes decreased with higher commuter travel frequency (*ρ*, 19% of changes in total epidemic sizes) and stronger distance weighting in the underlying gravity model (*ζ*, 15% of changes in total epidemic sizes). This suggests that these factors not only facilitate spatial disease spread but also determine whether outbreaks are larger in urban than in rural environments.

### Efficacy of different control strategies in changing disease landscapes

<u>Trace and isolation of all infected individuals</u> (trace all) was by far the most efficient control strategy in our simulations (**Fig 1**): no simulated scenario with ≥ 47% of infected individuals removed (κ) had a relative epidemic size > 5% of the respective baseline scenario. Lowering the epidemic size through isolation of infected individuals was less efficient for large transmission parameters (β, explaining 19% relative influence on changes in relative epidemic sizes, **Fig 2**).

**Fig 1.**
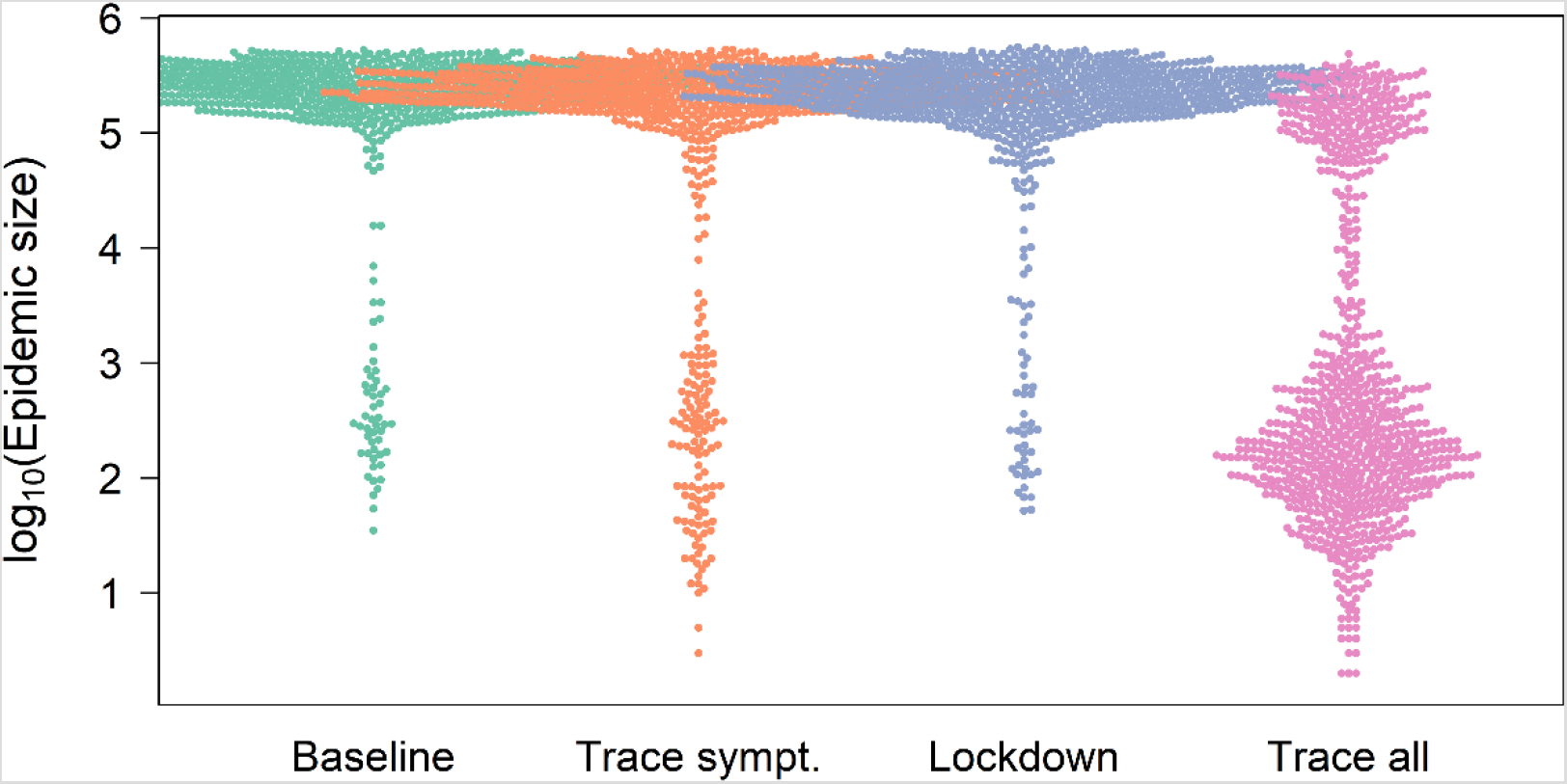
Distribution of the total COVID-19 epidemic sizes across an urban-rural gradient. Plot shows log_10_-scale epidemic size at metapopulation level resulting from simulating a large range of scenarios. Scenarios include: ‘Baseline’: no control strategy; ‘Trace sympt.’: isolation of a certain percentage of infectious/symptomatic virus carrier only; ‘Lockdown’: regional reduction of transmission parameters in response to a certain number of infectious/symptomatic virus carriers being present; ‘Trace all’: isolation of a certain percentage of infected individuals (i.e. those in the disease states exposed, asymptomatic virus carrier or infectious/symptomatic virus carrier). To aid visualisation, the plot is based on a random selection of 10,000 out of 40,000 simulation results.

**Fig 2.**
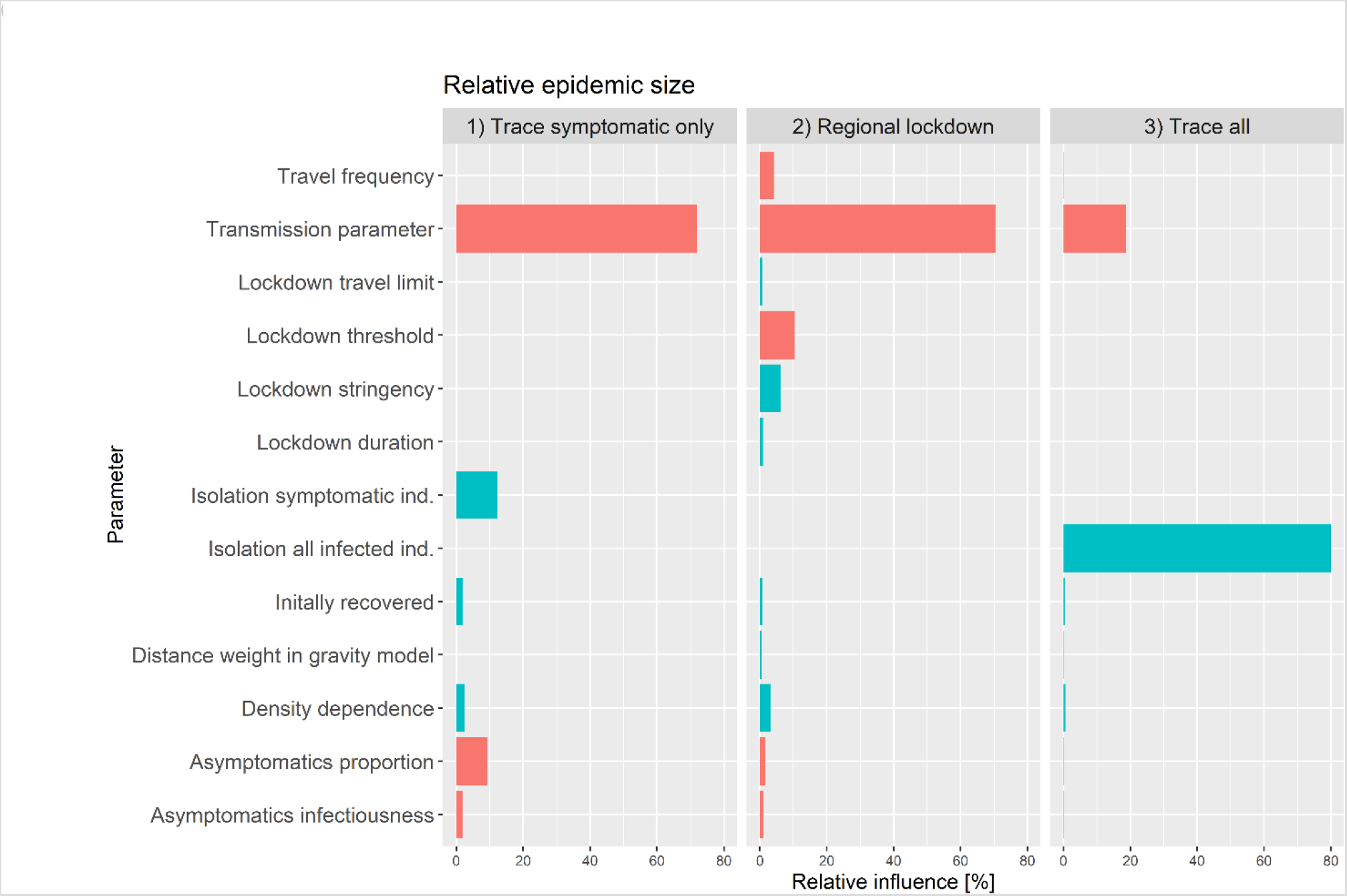
Relative influence of different parameters on the relative epidemic sizes. Relative epidemic sizes were calculated for simulations with three different control strategies compared to baseline scenarios of no COVID-19 control. The three different control strategies were ‘Trace symptomatic only’: isolation of a certain percentage of infectious/symptomatic virus carrier only; ‘Regional lockdown’: regional reduction of transmission rates in response to a certain number of infectious/symptomatic virus carriers being present; and ‘Trace all’: isolation of a certain percentage of individuals being infected in the disease states exposed, asymptomatic virus carrier or infectious/symptomatic virus carrier. Green bars indicate smaller and red bars larger relative epidemic sizes with increasing parameter values.

<u>Trace and isolation of symptomatic individuals</u> (trace symptomatic only) was of limited efficacy in lowering epidemic size in our simulations. The efficacy of these control strategies largely depends on small transmission parameters (β, 72% relative influence), whereas variation in the proportion of symptomatic individuals being isolated (ε) explained only 12% in relative epidemic sizes. The efficacy of this control strategy was further hampered by increasing proportions of asymptomatic cases (φ, 9% relative influence).

<u>Regional lockdown scenarios</u> appeared to be of limited efficacy in our simulations (**Fig 1**) and largely depend on small transmission parameters (β, 70% relative influence) (**Fig 2**). Their efficacy was sensitive to the regional threshold levels for lockdown implementation (α, 10% relative influence) and lockdown stringency (ϕ, 6% relative influence). A reduction of relative epidemic sizes to 5% of those of the respective baseline scenarios through regional lockdowns was only achieved for regional lockdown threshold levels of ≤ 1% the populations being symptomatic.

### Variation in control efficacy across urban-rural gradients

The strength of the urban-rural gradient in relative epidemic sizes resulting from <u>isolation of all infected individuals (E,A,I)</u> declined with increasing proportions of infected individuals isolated (κ, 46% relative influence, **Fig 3**) and increased with increasing transmission parameters (β, 24% relative influence), suggesting that larger transmission rates makes it relatively more challenging to control the spread in urban than in rural areas. In contrast, the more individuals are isolated (increasing κ), the more efficiently can epidemics be also contained in urban environments (i.e. resulting in less strong urban-rural gradients in relative epidemic size), despite a concentration of cases there, as depicted by mostly positive correlation coefficients of the urban-rural gradient in relative epidemic size (**Fig 4**).

**Fig 3.**
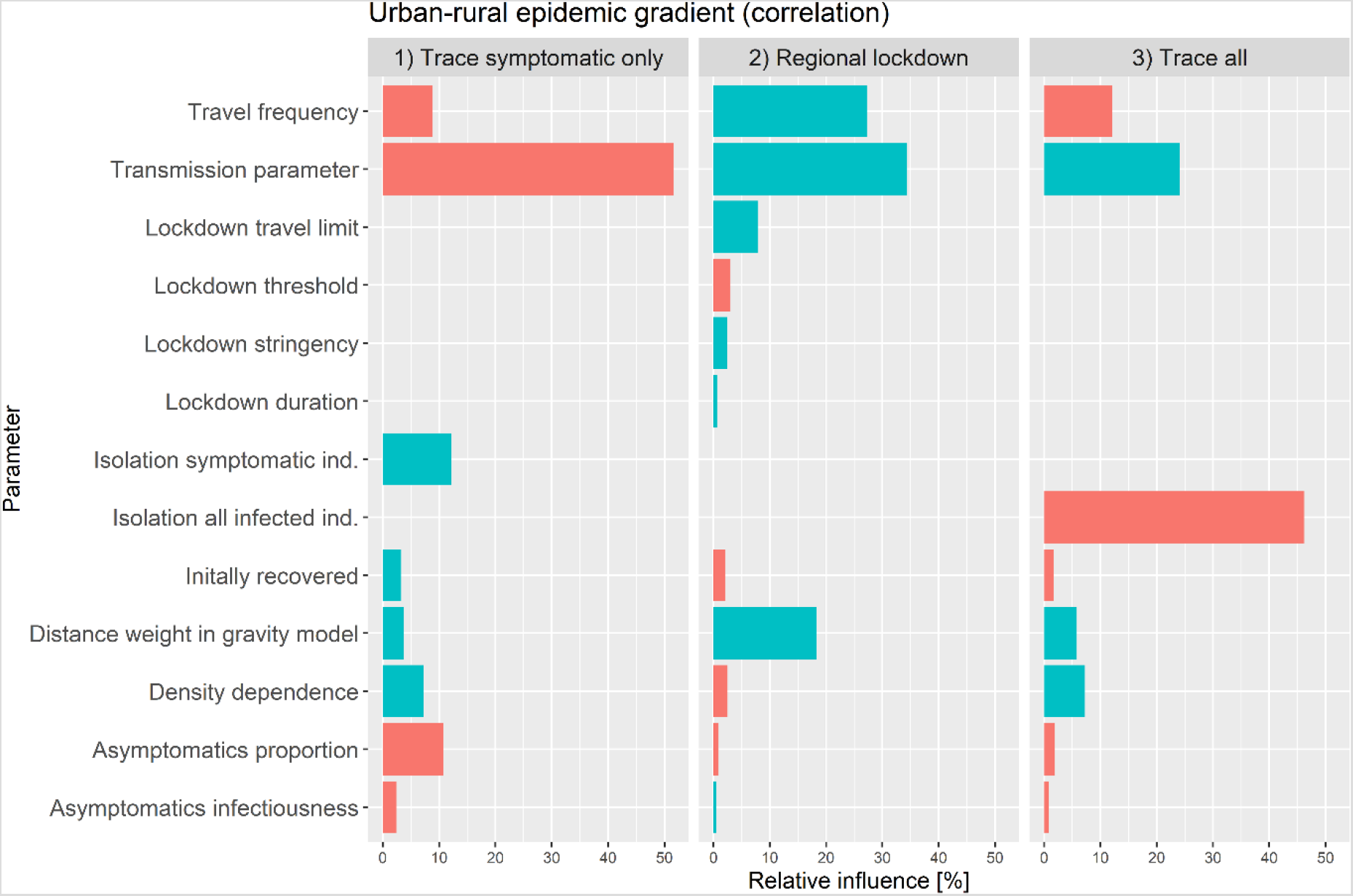
Relative influence of different parameters on the ‘urban-rural gradient’ (correlation coefficients of regional relative epidemic sizes with connectivity across all populations). Stronger correlations mean larger regional epidemic sizes in populations with increased connectivity, which are typically urban areas. Relative epidemic sizes were calculated for simulations with three different control strategies compared to baseline scenarios of no COVID-19 control. The three different control strategies were ‘Trace symptomatic only’: isolation of a certain percentage of infectious/symptomatic virus carrier only; ‘Regional lockdown’: regional reduction of transmission rates in response to a certain number of infectious/symptomatic virus carriers being present; and ‘Trace all’: isolation of a certain percentage of individuals being infected in the disease states exposed, asymptomatic virus carrier or infectious/symptomatic virus carrier. Green bars indicate decreases and red bars increases in correlation strength with increasing parameter values.

**Fig 4.**
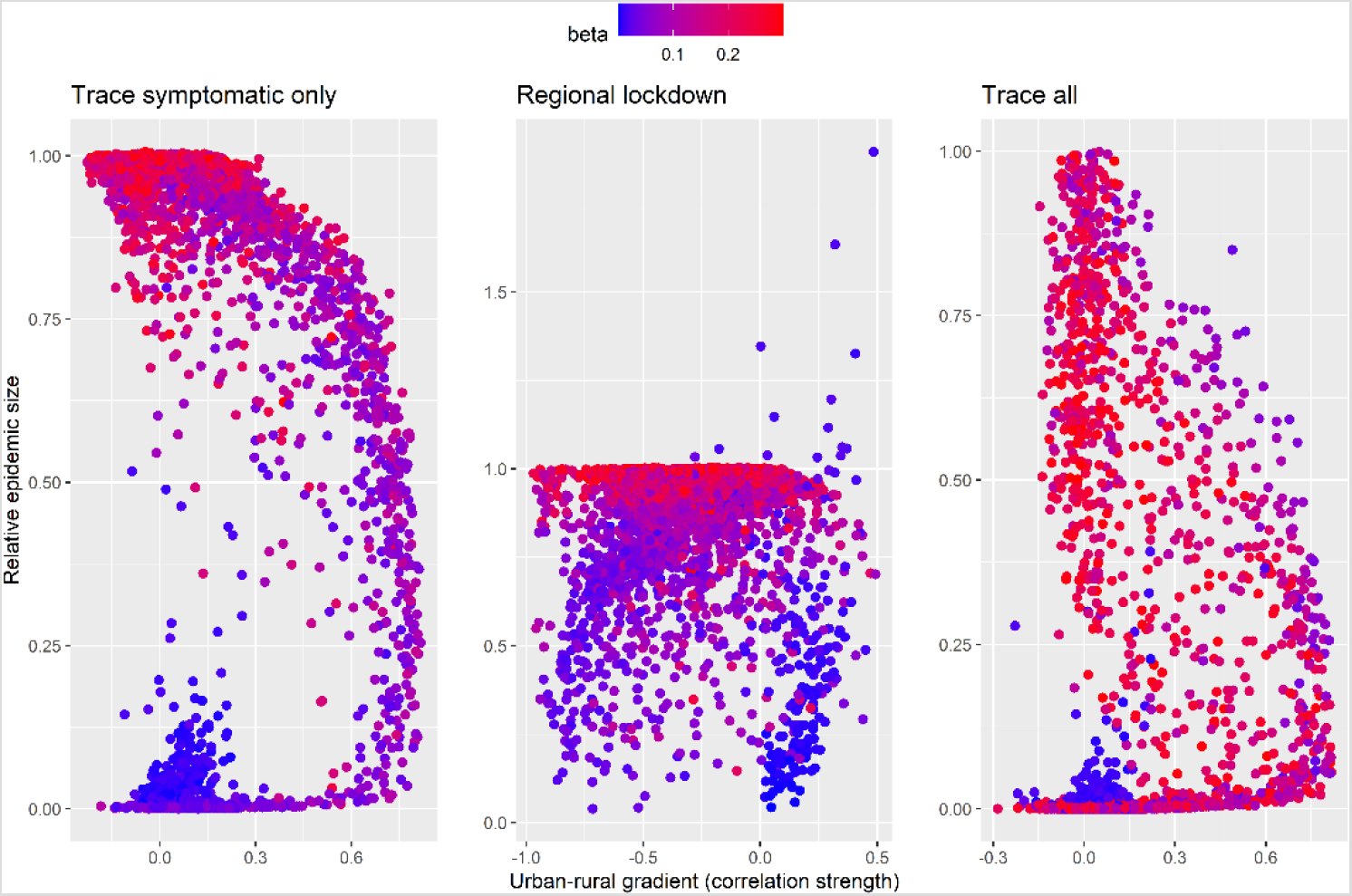
Relationship between overall relative epidemic size for different control measures and the underlying urban-rural gradient in epidemic size among populations. Relative epidemic sizes were calculated for simulations with three different control strategies compared to baseline scenarios of no COVID-19 control. The urban-rural gradient in epidemic size’ was computed as the strength of correlation the regional relative epidemic size and the respective population-level connectivity index. The three different control strategies were ‘Trace sympt.’: isolation of a certain percentage of infectious/symptomatic virus carrier only; ‘Lockdown’: regional reduction of transmission rates in response to a certain number of infectious/symptomatic virus carriers being present; ‘Trace all’: isolation of a certain percentage of individuals being infected in the disease states exposed, asymptomatic virus carrier or infectious/symptomatic virus carrier. Each point represents the outcome from a simulation with a different baseline scenario, coloured according to the respective value of transmission parameter (β).

The completely opposite effect was found for the *isolation of symptomatic individuals only (I)*. The strength of the urban-rural gradient in relative epidemic size declined with increasing transmission parameters (β, 52% relative influence) but increased with increasing proportions of symptomatic individuals isolated (ε, 12% relative influence). Hence, larger transmission rates make reduction in epidemic size by isolation of symptomatic individuals only more challenging in rural rather than in urban areas. The urban-rural gradient in relative epidemic size further decreased with larger proportions of asymptomatic cases (φ, 11% relative influence), decreased with higher commuter travel frequency (*ρ*, 8% relative influence) and increased with stronger density dependence in contact numbers (δ, 7% relative influence, **Fig 3**).

In response to <u>regional lockdown</u> strategies, the strength of the urban-rural gradient in relative epidemic size increased with increasing transmission parameters (β, 34% relative influence), increasing travel frequencies (27% relative influence), and stronger distance weighting in the underlying gravity model (*ζ*, 18% relative influence, **Fig 3**).

## Discussion

Decision-making to balance efficient COVID19 control with socio-economic pressures is a challenging task against the backdrop of asymptomatic disease spread and ever-changing disease landscapes. We show that isolation of symptomatic cases, or regional lockdowns in response to local outbreaks, have limited efficacy in terms of reducing overall epidemic sizes, unless overall transmission rate is kept persistently low. Isolation of non-symptomatic infected individuals, which may be detected by effective test and trace approaches, is pivotal to reduce overall epidemic size over a wider range of transmission scenarios. By considering an ‘urban-rural epidemic gradient’ as the strength of correlation between regional epidemic size and connectivity within a region, we show that under certain conditions, control measures are of limited efficacy in urban compared to rural areas. Intervention strategies focusing on the isolation of non-symptomatic individuals and regional lockdowns, for example, had the strongest urban-rural outbreak gradients at high transmission rates. In contrast, interventions targeting symptomatic virus carrier only had the reverse effect.

Our results emphasise the importance of efficient detection of infectious individuals through test and trace approaches for containing the spread of COVID-19 [2, 19, 20], while also uncovering that some methods will be less efficient in urban areas under the post-lockdown situation unless transmission rates are kept constantly low.

Efficient removal of all infectious individuals (including non-symptomatics) has the potential to restrain total epidemic size by successfully suppressing landscape-scale disease spread and the corresponding source-sink dynamics of how the disease may spread and re-emerge among populations. We found regional lockdowns to be only effective in terms of reducing overall epidemic size if implemented at low threshold levels and low transmission rates. This is likely due to the fact that only under these conditions can landscape-scale spread of the disease be avoided. These findings are in line with previous suggestions that temporary lockdowns do not necessarily contain overall epidemic size in a metapopulation context over medium to long time periods [21], even if they may be useful for reducing local case number over short time periods to avoid an overload of health capacities [22–24].

In practice, the prominent example of the locally restricted lockdown implemented in the city of Leicester in the UK, which began in June 2020 is just one example of mounting evidence that regional lockdowns do not necessarily see an reduction in disease transmission during the following weeks [25], which would ideally prevent spread of the virus beyond the local context. This slow response of incidence decline following regional lockdowns is in line with our finding and more general suggestions that disease with asymptomatic transmission pathways can only be controlled with intensive test and trace approaches [26].

Surprisingly, we found travel frequency and possible density dependence in contact frequency to have rather small relative impact on overall epidemic size compared to the transmission parameter (**Fig 2**). Despite the recognised importance of connectivity, travel patterns and metapopulation structure on disease spread [27–29] our results highlight the importance of overall transmission rates on disease spread and epidemic size. This has important management implications, as it points to measures that might allow for continuous long-term lowering of transmission rates. Such measures, we suggest, are considerably more efficient than any short-term measures of changing control stringency in response to actual case numbers for reducing the overall epidemic size.

We found the magnitude of transmission rate to also determine the success of different control strategies in urban versus rural areas, leading to varying urban-rural epidemic gradients in response to varying transmission rates and different control strategies (**Fig 3**). For interventions focused on isolating both non-symptomatic and symptomatic individuals and regional lockdowns, our results reveal the strongest urban-rural epidemic gradients at high transmission rates, indicating a reduced efficacy of such control measure in urban areas under these conditions. These results suggest that at high transmission rates, the urban-rural epidemic gradient is enforced by the overall poorly curbed disease spread at metapopulation level (see **Fig 4**). Conversely, we found the urban-rural gradient in epidemic sizes to be mostly masked at high transmission rates for measures targeted at symptomatics only, suggesting that that these measures (which are generally of moderate to low efficacy), would not contain disease spread at metapopulation level unless transmission rates are kept constantly low (see **Fig 4**). Exploring such effects warrants further investigation based on empirical data and relevant spatiotemporal models of disease spread under variable conditions of contact frequencies and control efforts. Such more detailed research may also account for first insights into variable compliance in response to intervention strategies. A recent study, for example, found slightly larger reductions in average mobility in high density than low density areas in the UK [30].

In contrast to many forensic COVID-19 models that have focused on forecasting real outbreaks in specific locations [1, 9, 10] our model is strategic, with a focus on exploring general mechanisms emerging from across a large range of modelled scenarios. A direct match to the ongoing epidemic in the study area is unfeasible because we do not account for any particular real-world starting conditions nor the temporary changes in human interactions in response to changing policy. Also, as we are not aware of detailed estimates of relevant epidemiological parameters such as how transmission rate varies among age groups in our study area, we do not account for age structure in our model, even though, as it has been shown, COVID-19 effects and expression of symptoms are rather different between children and adults [31]. These effects might be exacerbated by a potential systematic variation in demographic community composition in urban and rural areas. However, with an area-wide spread of COVID-19 in our study area and a concentration of cases in urban communities during the first six months of the epidemic, some general patterns found in model output and empirical data appear to be compatible (personal observations). Given more detailed data of spatiotemporal disease spread and better estimates of epidemiological key parameters, future studies may narrow down the currently intractable large parameter space through statistical approximation methods in order to identify when and how management efforts may results in disease extirpation versus long-term persistence [32].

The most important implication from our model is that priority should be given to any reliable and feasible measures that constantly keep transmission rate low as opposed to relying on local lockdowns to stamp out outbreaks. The success of any short-period interventions is limited if overall transmission rate remain high and facilitate disease spread within and among populations. We conclude that in the absence of an intervention strategy that would ensure rapid eradication of COVID-19, different intervention strategies do not work as efficiently in urban as in rural communities. Priority should thus be given to further research on how the most vulnerable individuals can be best protected at minimal cost for entire metapopulations. While post-lockdown situations of low transmission rates and reduced cases number are tempting to ease interventions, we believe that ongoing source-sink dynamics of disease spread cannot be ignored. Successful regional disease control during a pandemic should not ignore the fact that those communities that successfully escaped the first epidemic waves remain the most vulnerable because of large pools of individuals yet to be exposed to COVID-19.

## Data accessibility

The R code for this study can be found on GitHub https://github.com/konswells1/COVID-19-LSOA-metapopulation-model.

## Data Availability

The R code for this study can be found on GitHub https://github.com/konswells1/COVID19-LSOA-metapopulation-model.

## Acknowledgments

We acknowledge the support of funding from the Welsh Government for this project, and also the Supercomputing Wales project, which is part-funded by the European Regional Development Fund (ERDF) via the Welsh Government.

## Author contributions

KW – Conceptualization, Formal analysis, Writing – original draft, Writing – review & editing

ML – Conceptualization, Formal analysis, Writing – review & editing

BC – Formal analysis, Writing – review & editing

BL – Formal analysis, Writing – review & editing

RRK – Formal analysis, Writing – review & editing

ALL – Formal analysis, Writing – review & editing

SDWF – Formal analysis, Writing – review & editing

MBG – Conceptualization, Formal analysis, Funding acquisition, Writing – review & editing

